# Analysis of common glucocorticoid response genes in childhood acute lymphoblastic leukemia in vivo identifies cell cycle but not apoptosis genes

**DOI:** 10.1101/2020.07.11.20148890

**Authors:** Tatsiana Aneichyk, Stefan Schmidt, Daniel Bindreither, Armin Kroesbacher, Nikola S Mueller, Bernhard Meister, Roman Crazzolara, Georg Mann, Renate Panzer-Gruemayer, Reinhard Kofler, Johannes Rainer, Stephan Geley

## Abstract

Glucocorticoids (GCs) are an essential component of acute lymphoblastic leukemia (ALL) therapy. To identify genes mediating the anti-leukemic GC effects *in vivo*, we performed gene expression profiling of lymphoblasts from 46 children during the first 6-24h of systemic GC mono-therapy. Differential gene expression analysis across all patients revealed a considerable number of GC-regulated genes (190 induced, 179 repressed at 24h). However, when 4 leukemia subtypes (T-ALL, ETV6-RUNX1^+^, hyperdiploid, other preB-ALLs) were analyzed individually only 17 genes were regulated in all of them showing subtype-specificity of the transcriptional response. “Cell cycle-related” genes were down-regulated in the majority of patients, while no common changes in apoptosis genes could be identified. Surprisingly, none of the cell cycle or apoptosis genes correlated well with the reduction of peripheral blasts used as parameter for treatment response. These data suggest that (a) GC effects on cell cycle are independent of the cell death response and (b) GC-induced cell death cannot be explained by a single transcriptional pathway conserved in all subtypes. To unravel more complex, potentially novel pathways, we employed machine learning algorithms using an iterative elastic net approach, which identified gene expression signatures that correlated with the clinical response.

## Introduction

Synthetic glucocorticoids (GC) are essential components in the treatment of childhood acute lymphoblastic leukemia (chALL) but which chALL patients will respond to GC therapy is not known. In addition, GC-resistance is a poorly understood phenomenon that limits GC therapy [1]. According to the AIEOP-BMP protocol (http://www.bfm-international.org/aieop_index.html) patients undergo 1 week of systemic GC monotherapy, after which peripheral blood counts are included in the stratification of patients into high and standard/medium risk groups, because a poor GC-response, i.e., >1000 blasts/µl on day 8, is associated with a high risk of relapse [2]. The major subtypes T-ALL and precursor B-ALL (preB-ALL) are further subdivided according to immunophenotype or molecular markers, such as ploidy, chromosomal translocations, such as BCR-Abl, ETV6/RUNX1 in preB-ALL, mutation of NOTCH1 in T-ALL and others [1].

To improve the efficacy of GC treatment and to overcome primary and secondary GC-resistance [3-5] the molecular mode of action of GC *in vivo* needs to be better understood. GCs mainly act *via* the GC receptor (GR, *NR3C1*) through transcription regulation [6-8]. Upon ligand binding the cytosolic GR [9] is transported into the nucleus [10,11], where, upon binding to GC-response elements (GREs), the GR directly induces or represses transcription. The GR also collaborates with other transcription factors, such as AP1 [12], HES1 [13], STAT5 [14], NF-κB [15,16], and others [17], to regulate gene expression in the presence or absence of GREs. GCs can also exert non-genomic effects, which are, however, less well understood [18]. Because GCs activate a transcription factor, the anti-leukemic effects are likely to be initiated by changes in gene expression, although other mechanisms have also been proposed [19-21]. Analysis of leukemia cell lines suggested that the anti-leukemic effects of GCs are due to inhibition of proliferation and induction of cell death via regulating the expression of numerous genes [22-28], including some controlling apoptosis [29,30]. Whether regulation of these genes underlie the action of GCs *in vivo* in children with ALL is, however, not clear. To overcome this gap, we generated whole genome expression profiles from 46 children with ALL during GC monotherapy and correlated them, for the first time, with the clinical response. As detailed below, we observed GC treatment-associated regulation of mRNAs for key regulators of mitosis in all chALL subtypes but no consistent alteration in genes controlling cell death.

## Materials and Methods

### 1. Patients, lymphoblasts and expression profiling

Forty six children with ALL admitted to the Department of Pediatrics, Innsbruck Medical University (n=38), or the St Anna Children’s Hospital in Vienna (n=8) and treated according to BFM protocol 2000 (n=43) and AIEOP-BFM 2009 (n=3) were prospectively enrolled in this study, approved by the Ethics Committee of Innsbruck Medical University (EK1-1193-172/35) with written informed consent obtained from parents or custodians. The relevant clinical data are detailed in the Supplementary Table S1 and Figure 1.

**Figure 1:**
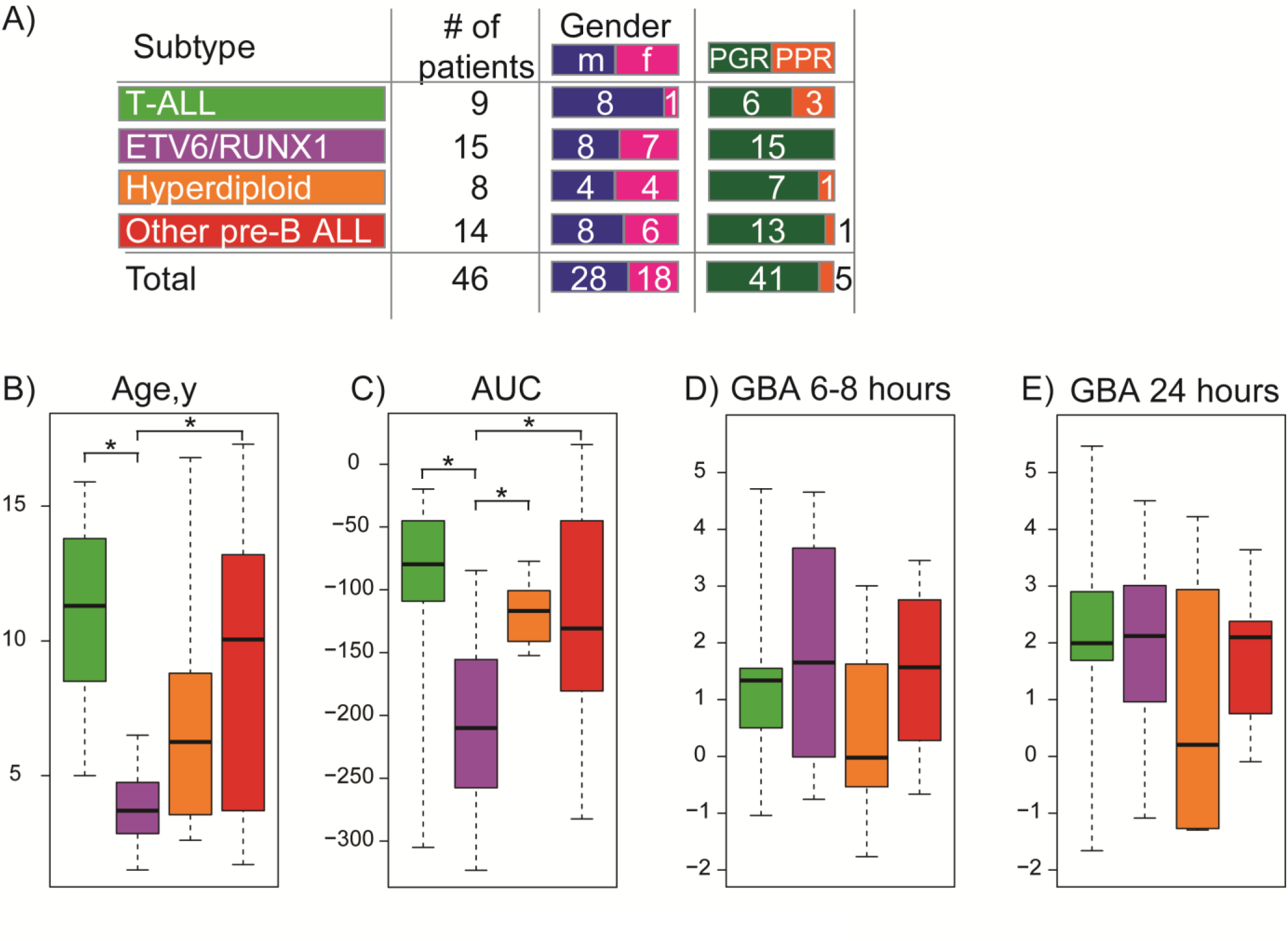
Summary of patient characteristics. The graphs summarize (A) the number of patients per subtype, sex, and prednisolone response (PGR, PPR: prednisolone good or poor responders), (B) the age distribution of the patients, (C) their blast response to systemic GC monotherapy (AUC, area under the curve – see Material and Methods), (D) and (E) the change in serum levels of active GC after 6-8h (D) and 24h (E) GC treatment as measured by GC bioactivity assay (GBA). Shown are box plots with mean values, and whiskers of 25% from all patients per subtype (color code as in panel A) where corresponding data were available (see Supplementary Table S1 for individual data). Significance values (p<0.05) between groups are indicated by an asterisk.

Blood samples were taken from patients prior to, and 6-8 and 24h after the first intravenous application of prednisolone at 20mg/m^2^ at 3 doses/day. Blasts were isolated to >90% purity and total RNA subjected to expression profiling using Affymetrix U133 Plus 2.0 microarrays. Blast counts were recorded prior to and daily during treatment. GC bioactivity in the blood at the time of sample recovery was determined as published [31]. Details regarding blood sampling, treatment protocol, lymphoblast purification, RNA preparation and Affymetrix gene chip analysis including all quality control measures have been published previously [32].

### 2. Data preprocessing

For microarray raw data preprocessing see Supplementary Information. Raw and preprocessed expression data have been deposited at the Gene Expression Omnibus (Accession number: GSE73578); *during the review process data are accessible using the link: http://www.ncbi.nlm.nih.gov/geo/query/acc.cgi?token=sfktcoyghzavrup&acc=GSE73578*.

### 3. Patients ALL subtype classification and GC-response quantification

Patients were assigned to 4 groups (Figure 1): T-ALL, ETV6/RUNX1 positive preB-ALL, hyperdiploid preB-ALL, and a mixed group of preB-ALL (“other preB-ALL”). A detailed description of signature generation and patient classification can be found in Bindreither et al. [33] and in Supplementary Table S1. The response of the patients to GC treatment was evaluated by calculating the area under the curve (AUC) from the blast counts during the first 72h of treatment (Supplementary Figure S1).

### 4. Differential gene expression and GO analyses

M-values (log2 fold change values) representing differential gene expression between the 6-8 or 24h time point and the time point prior to treatment initiation were obtained employing Bioconductor’s *limma* package [34] and p-values adjusted for multiple hypotheses testing according to Benjamini and Hochberg (BH) [35]. Genes with an absolute M-value larger than 0.7 (i.e. > 1.6-fold regulation) at a 5% false discovery rate (i.e. adjusted p-value < 0.05) were considered as significantly differentially expressed.

Gene ontology analyses were performed using R package topGO with algorithm “weight01” and Fishers statistics according to Alexa and Rahnenfeld (https://www.bioconductor.org/packages/3.3/bioc/html/topGO.html).

### 5. Regression models

Two different regression models were used to evaluate the relationship between gene expression/regulation and the response to GC-treatment as measured by AUC after 72h (*AUC*): “simple regression”, modeling gene expression/ regulation as dependent only on *AUC*, and “extended regression”, incorporating time point, ALL subtype and gender as adjustment factors. The detailed description of regression models are provided in Supplementary Information. The p-values for the *AUC* coefficients were adjusted for multiple hypotheses testing [35].

### 6. Elastic-net regression

For elastic-net regression, gene expression was adjusted for subtype, gender and time point by fitting a regression model and among the regulated genes, we focused on likely direct targets due to the presence of GR-binding sites determined in chALL cell lines [36]. Elastic net regression was applied to the combined data set of adjusted average expression values and average regulation values of approximately 1000 probe sets, each using the R package glmnet [37] with the elastic net mixing parameter α set to 0.1 and the regularization parameter *λ* within 1 standard error from the minimum of the cross-validation error curve in the direction of increased regularization (see Supplementary Information for details).

## Results

### 1. Characterization of the transcriptional response to GC in lymphoblasts from children with ALL

In order to define the molecular mechanism underlying the antileukemic GC effects, we analyzed 46 chALL patients undergoing GC treatment. Clinical data, such as leukemia subtype, age distribution, clinical response and active GC serum levels of the patients are summarized in Supplemental Table S1 and Figure 1. Microarray-derived expression profiles from purified leukemic blasts of these patients prior to, 6-8h and 24h after initiation of GC treatment were subjected to differential gene expression analysis. Using a false discovery rate (FDR) <0.05 and absolute log2 fold change > 0.7 (M values), we defined 206 and 516 “GC-regulated” probe sets after 6-8h and 24h, respectively, with similar numbers of up versus down-regulated probe sets (Figure 2A, “all patients”). As detailed in Supplementary Table S2A, many of the corresponding genes contained GR-binding sites in their regulatory regions and have been found regulated in cell lines exposed to GC [36]. During the time of treatment, gene regulation was monotonic as we did not observe any gene with apparent “counter regulation” (significant induction after 6-8h followed by repression after 24h or *vice versa*). The mean M values were moderate, i.e., between -1.6 and +1.9 after 6-8h (Figure 2B) and - 2.1 and +2.3 after 24h (Supplementary Figure S2), and even on an individual patient level, regulations rarely exceeded 4-8 fold changes (Supplementary Table S2B). Thus, GC-treatment affected many genes (144 after 6-8h and 340 after 24h) but, in most cases, by only mildly changing their expression.

**Figure 2:**
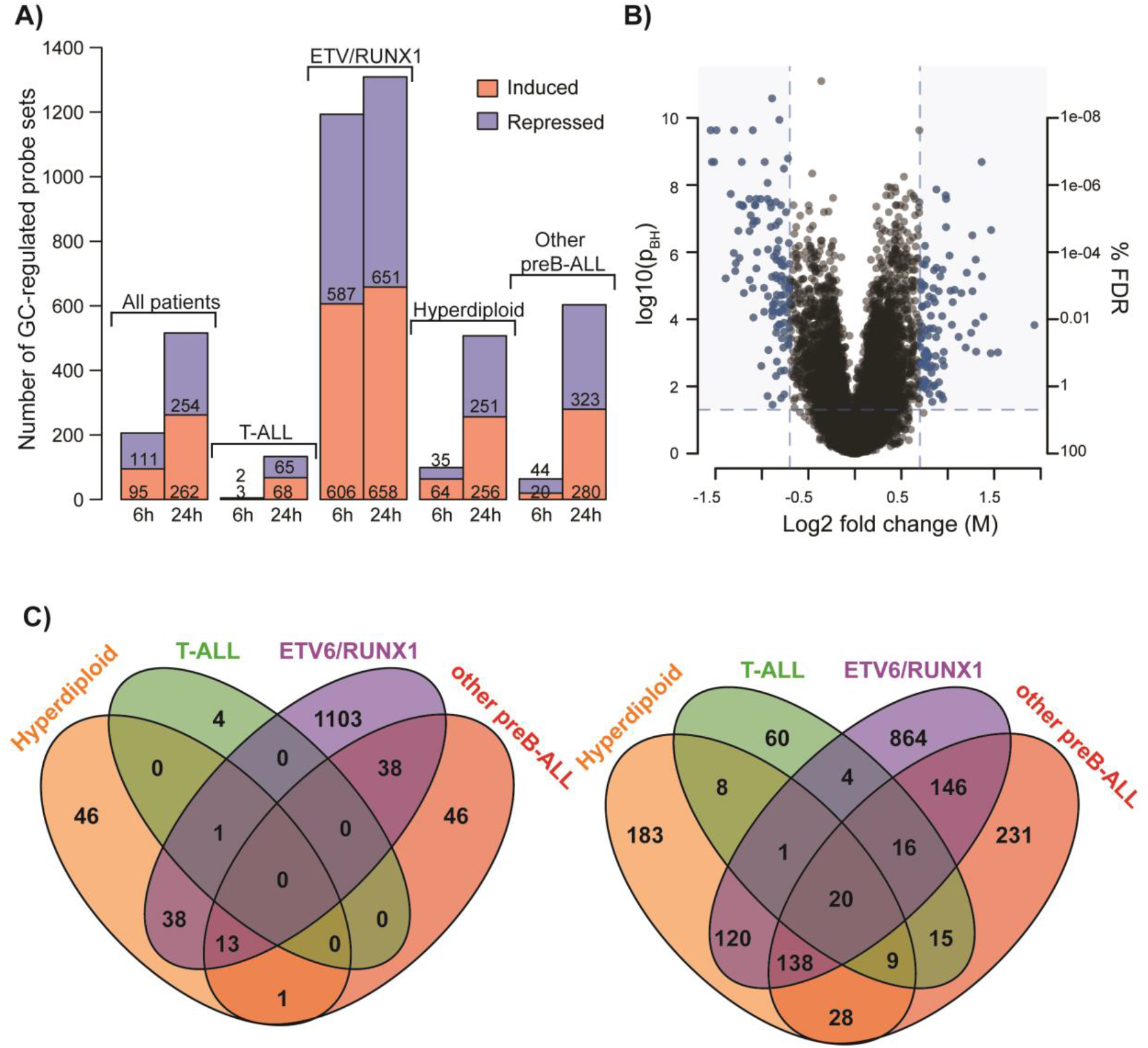
The transcriptional *in vivo* response to GC. (A) Panel A shows the number of GC-regulated probe sets after 6-8h and 24h GC monotherapy in all 46 patients and in the subgroups (all probe sets in Supplemental Table S2). (B) A “Volcano plot” showing the mean M values of all 46 patients GC-treated for 6-8h (M) and the BH adjusted p values (left scale in log10) or percent false discovery rate (right scale in log2). (C) Venn diagrams showing number of significantly regulated probe sets in the 4 subgroups after 6-8h (left panel) or 24h (right panel).

To address whether the transcriptional response was conserved between the various ALL subtypes, we performed differential expression analysis for every ALL subtype. As can be seen in Figure 2A, the molecular subtypes showed dramatic differences in their response to GC and not a single probe set was significantly regulated in all 4 subtypes after 6-8h and only 20 probe sets (17 genes) after 24h (Figure 2C). Although this underestimates the extent of common GC-regulated genes due to the arbitrarily set cut-off for M-values (many genes revealed regulation in several or all subtypes but just failed to reach a mean M-value of >0.7 and/or a pBH of <0.05), several genes were significantly regulated in only one molecular subtype but not regulated in others (as defined by a mean M-value ≤0.2 in any of the probe sets corresponding to a given gene, Supplementary Table S2A). Moreover, several genes were significantly induced in some but repressed in other subtypes (designated “both” in Supplementary Table S2A). Taken together, *in vivo*, the GCs activate a large number of genes in chALL blasts but to a relatively moderate extent, that increases over time, and comprises both a small number of common and many more ALL subtype-specific genes.

### 2. Assigning pathways to GC-regulated genes in leukemia treatment

Next, we performed gene ontology (GO)-term analyses for the genes found to be significantly regulated either at 6-8h or 24h (206/144 and 516/340 probe sets/genes, respectively, see Supplementary Table S2A). This GO term analyses revealed a dramatic enrichment of cell cycle-related terms at both time points, i.e., of the 43 GO terms significantly associated with these genes, 33 fell into the category “cell cycle” (see Supplementary Table S3 for details). Interestingly, at the same level of stringency, GO terms related to apoptosis or cell death induction did not reach significance levels suggesting that, unlike GC-induced cell cycle arrest, GC-induced cell death is not associated with a set of known cell death-inducing genes.

Since an individual gene responsible for cell death induction would not be detected by GO term analyses, we manually scrutinized all GC-regulated genes (mM>|0.7|; pBH<0.05) for literature-based evidence for a direct function in cell death regulation. This analysis revealed induction of the caspase-3 substrate DFNA5 [38] (induced at 6-8h and 24h) and the pro-apoptotic BH3-only molecules BMF and BCL2L11/BIM (both induced at 24h) but also repression of the pro-apoptotic BH3-only protein HRK (6-8 and 24h) and induction of the pro-survival BCL2 family member BCL2A1 (24h).

### 3. Identification of potential effectors of the blast response in vivo

To delineate whether GC-regulated genes correlate with the clinical response, we determined the area under the curve (AUC) change in peripheral blood blast counts during the first 72h of GC treatment. The AUC was selected as a more homogenous and robust parameter for correlation analysis than blast counts at a certain time point because it is derived from several measurements over 72h and thus reflects the dynamics of blast numbers and is less likely to be influenced by measurement errors (for examples see Supplementary Figure S1).

First, we performed hierarchical clustering of the transcriptional response of each patient after 6-8h (Figure 3) and 24h (Supplementary Figure S3). To reduce the number of genes entering this analysis we concentrated on potentially direct targets of the GR, thus analyzing 49 genes that were GC-regulated at 6-8h, and 110 genes regulated at 24h and contained potential GR binding sites, as determined previously using ChIP-chip data identifying GR binding sites (GBS) in childhood T-ALL (CCRF-CEM-C7H2) and/or preB-ALL (NALM6) cell lines [36]. This analysis revealed 2 main clusters of patients (Figure 3 and Supplementary Figure S3 for 6-8 and 24h, respectively). All T-ALL children fell into cluster I at both time points, while ETV6/RUNX1 leukemia cases were found in cluster II (all at 24h, all but 1 at 6-8h). The children from the hyperdiploid and “other” preB-ALL groups did not cluster, consistent with their more heterogeneous genotype. Moreover, there was no clustering with the serum GC levels (GBA) even though cluster II clearly showed a stronger transcriptional response. Importantly, although the overall blast response reflected in the AUC was weaker in cluster I compared to cluster II (6-8h: I: -98.3±70; II: -183.1±74.1; 24h: I: -101.1±70.3; II: - 166.5±80), there was no clear clustering with the blast response. For example, T-ALL case 61-KKI, a strong blast responder, clustered with the other (much weaker responding) T-ALLs in cluster I but not with the other good responders in cluster II suggesting that the transcriptional response is dominated by subtype specific genes rather than cell death inducing ones.

**Figure 3:**
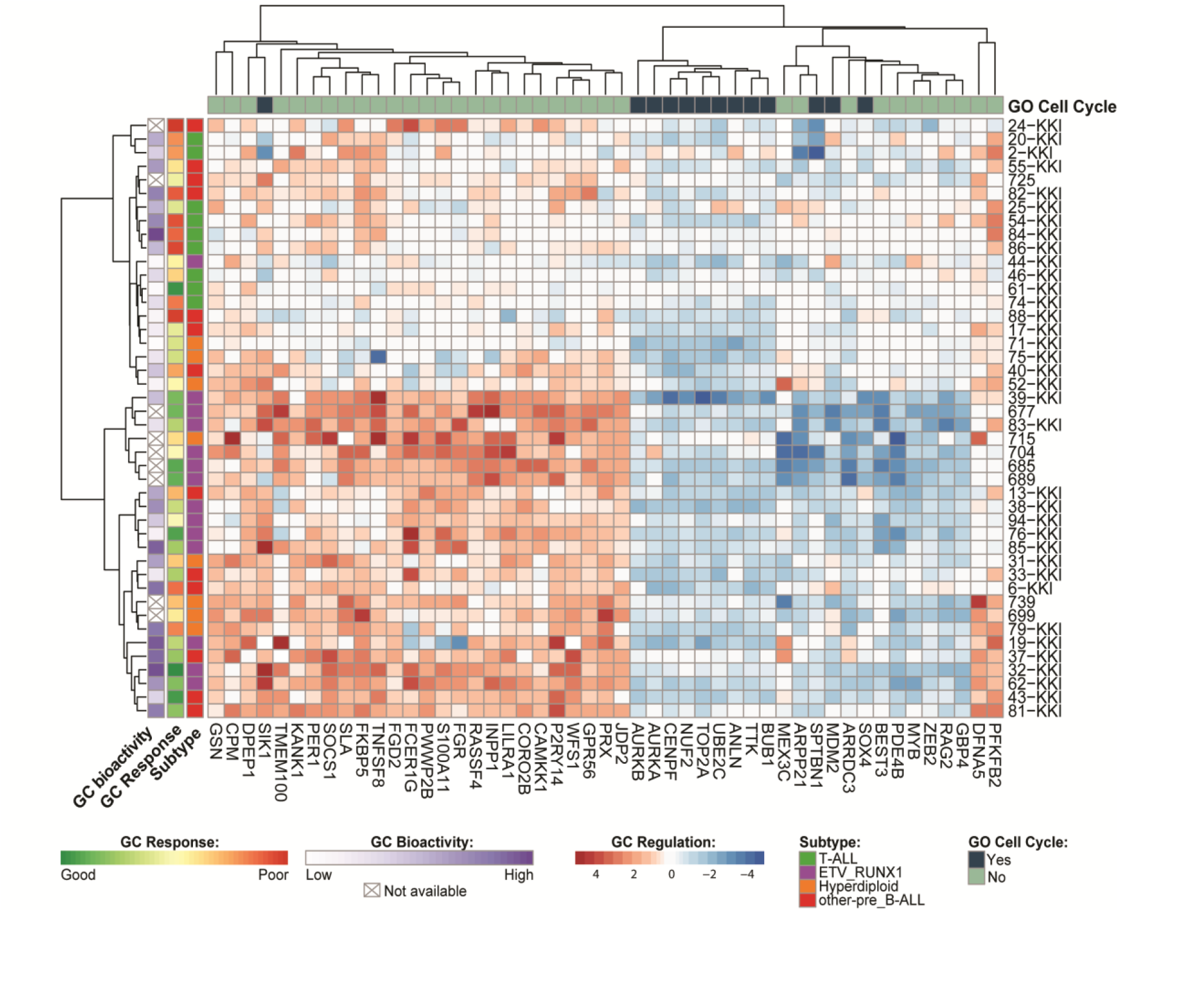
Hierarchical clustering of ‘direct’ GC response genes. “Heatmap” depicting 49 genes that contain a glucocorticoid receptor binding site and are regulated after 6-8h of systemic GC monotherapy. A corresponding heatmap for 24h is shown in Supplementary Figure S3, the values for the individual genes are summarized in Supplementary Table S2A.

The GC-regulated genes clustered in two main groups, i.e., genes mainly up- or down-regulated by GCs. Among 13 down-regulated genes, 9 were linked to the cell cycle as determined by GO-term analysis. None of these genes, however, correlated with the ’blast responses’. The lack of correlation between the regulation of cell cycle genes and clinical outcome suggests that GCs have separate effects on cell cycle regulation and cell death induction, as we have proposed for the CCRF-CEM chALL cell line model [39]. Among the non-cell cycle related genes, we found induction of *JDP2 [40], P2RY14 [41], RASSF4 [42], CD30 [43], SOCS1 [44], PER1 [45], KANK1 [46] and PFKFB2 [47]*, as well as suppression of *PDE4B [48], SOX4 [49] and MDM2 [50]*, which have been shown to control proliferation or cell survival in leukemia or other malignancies.

Next, we correlated the blast response (AUC) with gene regulation at 6-8h and 24h of all probe sets using ordinary least squares regression (“simple regression”). At 6-8h, no gene reached significance and the best coefficient of correlation (R^2^) was 0.41 (data not shown). After 24h, 7 genes reached significance but their R^2^ values were still moderate (0.45 or less). Moreover, 3 of these genes (*RTN1, PLEKHA1*, and unassigned 237520_x_at) were regulated only weakly and were not among the 2307 GC-regulated probe sets defined as having a mM<0.7; the remaining 4 were regulated only in patients of the *ETV6/RUNX1* subtype (*BEST3, PLEK, VSIG10*) or in “others” (*S100A4*) but not in all patients. Hence, none was a good candidate explaining the ’blast response’ in all subtypes. Importantly, the above described GC-induced apoptosis related genes *BMF, BCL2L11/*BIM and DFNA5/*GSDME* showed no correlation with the ’blast response’, making them unlikely candidates for common cell death inducers by GCs in chALL patients.

Inclusion of additional factors to the model (“extended linear regression”), such as time of measurement of the blast response, subtype and/or gender, improved the coefficient of determination, *R*^2^, to at most 0.69, but none of the genes with significant p-value for the response coefficient (30 genes with p-values from 9.7e-05 to 0.05) had *R*^2^ higher than 0.47 (Table 1 and Supplementary Table S4). Given the above, this analysis suggests that there is no single GC-regulated gene that could explain the blast response in these patients. One explanation for the failure to detect a conserved cell death pathway in our chALL cohort might be that the pathway underlying the blast response depends on the cellular context, a hypothesis that is addressed in the next section.

**Table 1:** Genes detected as potential effectors of GC-response by regression analysis. A single probe set per gene is selected with the lowest p-value for the AUC coefficient. The complete data set, with all genes, coefficients and p-values can be found in Supplementary Table S4.

| Gene | pAUC | R<sup>2</sup> | Gene | pAUC | R<sup>2</sup> |
| --- | --- | --- | --- | --- | --- |
| FAM19A4 | 9,70E-05 | 0,40 | WDFY1 | 0,0298 | 0,26 |
| CCDC69 | 2,30E-03 | 0,47 | ELL3 | 0,0400 | 0,39 |
| ANTXR1 | 5,58E-03 | 0,33 | IQCK | 0,0400 | 0,41 |
| LRRC34 | 7,12E-03 | 0,34 | ERBB4 | 0,0400 | 0,27 |
| LOC101060019 | 7,12E-03 | 0,36 | CPEB4 | 0,0400 | 0,36 |
| CRISP2 | 7,12E-03 | 0,32 | SLC26A3 | 0,0400 | 0,22 |
| CALCOCO2 | 0,0137 | 0,41 | LPCAT4 | 0,0416 | 0,23 |
| ESRP1 | 0,0138 | 0,31 | SERPINA1 | 0,0416 | 0,31 |
| NPAS2 | 0,0154 | 0,30 | C6orf10 | 0,0423 | 0,24 |
| KCNK1 | 0,0166 | 0,30 | INVS | 0,0437 | 0,24 |
| ERMP1 | 0,0166 | 0,40 | CALML5 | 0,0464 | 0,29 |
| RHBDL3 | 0,0180 | 0,24 | RP11-26J3.1 | 0,0464 | 0,28 |
| LOC153546 | 0,0199 | 0,24 | RP11-159D8.2 | 0,0464 | 0,27 |
| PTCRA | 0,0211 | 0,29 | LOC100506172 | 0,0464 | 0,27 |
| PRG4 | 0,0298 | 0,29 | SLMAP | 0,0495 | 0,25 |

### 3. Identification of potential modulators of the blast response in vivo

Apart from “effector” genes (genes that are regulated by GC) able to control cell death, other genes might influence the blast response even if they are not regulated by GCs (“modulator” genes). These “modulator” genes may act either as facilitators or as inhibitors and, hence, their expression level might correlate positively or negatively with the blast response (as measured by AUC).

To identify such modulator genes, we used ordinary least squares regression, with additional adjustments for chALL subtype, gender and the time of measurement of blast counts (Supplementary Table S5). For 232 probe sets representing 146 genes this model could explain over 80% of variability, indicating a much better performance than the previous model based on GC-regulations. In addition, among them we were able to identify 64 probe sets representing 42 genes with significant relationship between gene expression and blast response (see Table 2 and Supplementary Table S5). Altogether, this can serve as evidence that these genes might be potential facilitators or inhibitors of the GC response in patients, thereby contributing to the particular cell context that is necessary for the treatment response.

**Table 2:** Genes detected as potential modulators of GC-response by regression analysis. A single probe set per gene is selected with the lowest p-value for the AUC coefficient. The complete data set, with all genes, coefficients and p-values can be found in Supplementary Table S5.

| Gene | pAUC | R<sup>2</sup> | Gene | pAUC | R<sup>2</sup> |
| --- | --- | --- | --- | --- | --- |
| CD24 | 2,78E-10 | 0,92 | LILRA2 | 0,013 | 0,84 |
| CD79A | 1,95E-06 | 0,84 | INSR | 0,016 | 0,81 |
| ZNF296 | 9,73E-06 | 0,85 | CSRP2 | 0,019 | 0,83 |
| LRP10 | 3,29E-05 | 0,82 | LOC284757 | 0,019 | 0,89 |
| CTNNA1 | 5,15E-05 | 0,87 | ZFY | 0,022 | 0,95 |
| CIITA | 1,27E-04 | 0,86 | UTY | 0,022 | 0,93 |
| BIVM | 1,48E-04 | 0,85 | PSD3 | 0,024 | 0,91 |
| GRAP2 | 1,42E-03 | 0,82 | TFDP2 | 0,025 | 0,83 |
| CYTH3 | 1,55E-03 | 0,86 | IGF2BP1 | 0,026 | 0,83 |
| KCNN1 | 1,59E-03 | 0,91 | DSC2 | 0,028 | 0,95 |
| GPR132 | 1,77E-03 | 0,89 | WNK3 | 0,031 | 0,85 |
| ENPP4 | 2,74E-03 | 0,80 | ASL | 0,031 | 0,85 |
| POU2AF1 | 3,01E-03 | 0,83 | SAMD13 | 0,033 | 0,94 |
| CLIC5 | 3,50E-03 | 0,86 | GNG7 | 0,039 | 0,84 |
| EBF1 | 4,29E-03 | 0,92 | MAPK13 | 0,039 | 0,82 |
| SH2D1A | 4,52E-03 | 0,93 | XIST | 0,040 | 0,81 |
| FAM49A | 6,15E-03 | 0,82 | PRKCQ-AS1 | 0,042 | 0,83 |
| TSHR | 6,50E-03 | 0,81 | MYOCD | 0,048 | 0,94 |
| DSC3 | 6,84E-03 | 0,90 | UNC93B1 | 0,049 | 0,82 |
| NUCB2 | 8,22E-03 | 0,88 | ZCCHC7 | 0,049 | 0,90 |
| HLA-DOA | 0,011 | 0,88 | ATP8A2 | 0,049 | 0,86 |

### 4. Correlating “regulators”, “modulators” and the blast response in vivo

By definition, modulators on their own cannot cause GC-dependent effects but require GC regulation of effector(s) (also referred to as ’regulators’). To identify combinations of modulators and regulators that may cooperate, we implemented an elastic net regularization-based approach. As a result, we identified 98 candidate genes that in combination might explain the blast response in the patients (62 regulators and 36 modulators, Figure 4 and Supplementary Table S6). Using hierarchical clustering, we found that these genes form 4 clusters (gene sets): GC-repressed regulators (gene set 1), GC-induced regulators (gene set 3), modulators acting as inhibitors (gene set 2) and modulators acting as facilitators (gene set 4). Children who failed to regulate gene sets 1 and 3, and have an increased expression of inhibitors and reduced expression of facilitators form the cluster of worst responding patients (patient cluster 6 in Figure 4). Children regulating sets 1 and 3 (patient clusters 1, 3 and 4) or set 3 alone (patient cluster 2) comprised the better responding patients, with modulator gene expression apparently influencing the extent of the blast response. GO-term analyses of the genes identified as regulators or modulators did, however, not reveal an obvious pathway that may help to understand the clinical response to GCs. Nevertheless, based on the clear clustering results, the expression levels or regulations of the genes shown in Figure 4 appear to correlate with the clinical response.

**Figure 4:**
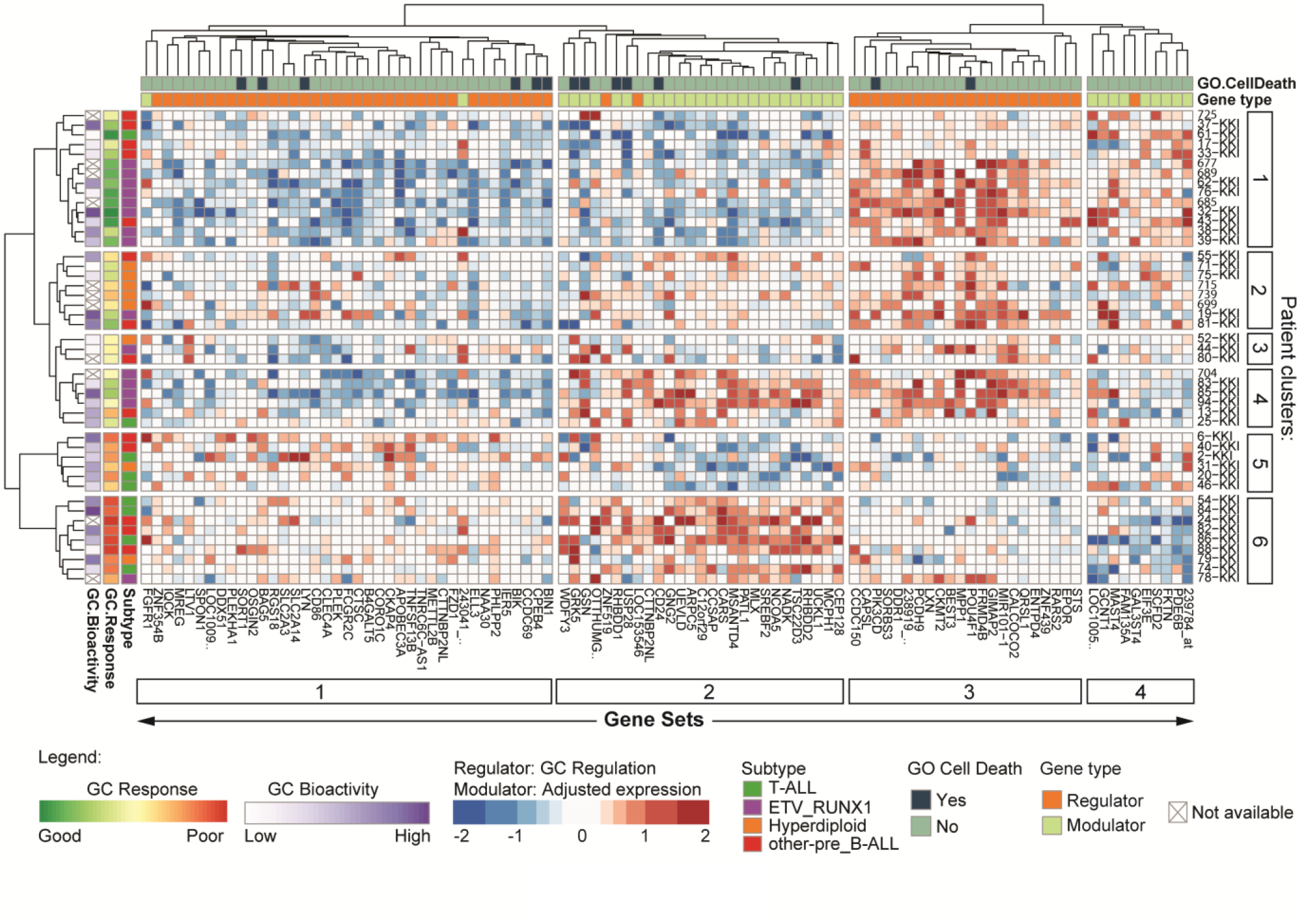
Elastic Net gene selection results. Gene is marked as “Regulator” and “Modulator” if gene regulation or gene expression were found to contribute to the response (AUC), respectively. For regulators, the color represents the average regulation in a patient from 0 to 6-8h and from 0 to 24h. For modulators, the color represents average adjusted gene expression in a patient (see Materials and Methods). GC response was measured as AUC in the first 72h of treatment.

## Discussion

Glucocorticoids induce cell death in cells of the lymphoid lineage and several genes have been suggested to provide a mechanistic explanation of GC-induced cell death [4,51,52]. These data were obtained from studying cell lines, patient samples or murine animal models but no canonical GC-induced cell death pathway in leukemia able to explain the phenomena of GC sensitivity and resistance could be extracted yet. This lack of a consensus pathway might be due to the fact that these experimental systems are very diverse, e.g., cell lines correspond to a single clinical case only and, in addition, do not necessarily represent the underlying disease as they may diverge from the clinical situation by mutations and adaptations acquired during the *in vitro* cultivation process. Similar restrictions apply to *ex vivo* treated primary samples, cells used in xenographs or other model systems. For example, GILZ (*TSC22D3*), a well-known GR target gene [53], was not identified to be strongly GC regulated in our patients because GILZ is already strongly expressed in the patients due to the action of endogenous corticoid hormones. When patient samples are then removed from the patient and kept in standard media, GILZ mRNA expression values drop dramatically but are rapidly re-induced by *in vitro* re-exposure to GCs (data not shown).

Alternatively, a consensus pathway might not exist, because GC effects are pleiotropic and may reduce the threshold of cell death induction, e.g., by dampening survival signals, or induce (untimely) differentiation, or interfere with metabolism, rather than inducing a cell death gene or pathway.

In this study, we determined gene regulation by GC *in vivo* in children suffering from ALL and, thus, provide a data set that is as close as possible to the *in vivo*, i.e., the clinical, situation. We first determined which genes were regulated by GC and correlated gene expression levels with clinical GC-effects (blast counts). Since we aimed at correlating the clinical response, as measured by peripheral blast counts, with gene expression changes during the first 24h, we used peripheral blood counts up to 72h, rather than taking the “classical” 8d time point used for stratification purposes or even later time points. Taking the AUC of blast counts over several time points (before treatment and at several time points up to 72h) was considered more robust than using a single time point because it included several measurements and reflected the dynamic changes during therapy. Moreover, in preliminary experiments the 48h and 72h time points gave similar results. Thus, given the above considerations we calculated a normalized AUC to adjust for different initial blast counts as read out for the clinical response. To determine GC-regulated genes, we used a modest M-value (log2 fold change) cut-off at 0.7, which corresponds to 1.62-fold change in gene expression compared to pretreatment levels. GCs influence the expression of several thousands of genes directly and indirectly with various degrees of magnitude of regulation. However, for correlation analysis between gene expression and clinical outcome, we used the whole data set without an M-value cut off to also test weakly regulated genes for their potential role in GC effects *in vivo*. The data set generated in this study is unique because it contains *in vivo* gene expression data from a considerable number (46) of children with ALL during the first 24h of systemic GC monotherapy. Each individual expression profile is linked to clinical data from an individual patient, including peripheral blood GC levels and peripheral blood blast counts, which provides a unique resource for correlation analysis between gene expression and regulation and the clinical response. Furthermore, this data set is a resource to validate GC-regulated candidate genes, identified in any model system for their potential role *in vivo*.

We have exploited this data set to analyze the *in vivo* mRNA response to GC in chALL cells and to correlate *in vivo* gene expression data with the clinical response to GC. Our study led to three important conclusions: 1. Among patients from at least 4 different types of ALL, GCs regulate only a limited number of common genes; 2. GCs regulate a large set of cell cycle genes but the extent of regulation did not correlate with a drop in blast counts; and 3. GCs do not induce a common set of apoptosis genes, suggesting that induction of cell death is highly cell type specific.

In our opinion, the number of patients (46) is sufficient to answer the general question of whether the anti-leukemic effects of GC (i.e., cell cycle arrest or cell death) are correlated with, and perhaps caused by, common changes in the gene expression profile shared in all types of childhood ALL. In fact, we observed repression of mRNA of cell cycle associated genes throughout all subtypes. Even though, for technical reasons, we don’t know how many and which cells in a given patient show this response, the fact that we can extract this response shows that shared transcriptional responses can be observed in a heterogeneous data set consisting of transcriptional responses of different populations of malignant lymphoblasts (T-ALL, different types of Pre-B-ALL) investigated in this study.

For this reason, we find it remarkable that not a single gene previously suggested to be involved in GC-induced apoptosis could be identified as associated with the blast response in this study. We concluded, therefore, that there is no common transcriptional response of cell death genes associated with GC-induced cell death in childhood ALL. Moreover, the same result, i.e., suppression of cell cycle-associated genes, but no shared cell death mRNA response, was derived when T-ALL (9) and pre-B-ALL (37) patients were investigated separately. Even the most homogeneous subgroup, i.e., ETV6/RUNX1 with 15 patients, produced the same results, confirming our conclusion. Similar findings, i.e., very heterogeneous GC responses and lack of a consensus cell death initiation pathway, were recently obtained in a study that compared GC-regulated genes in 10 B-ALL cell lines, 3 *ex vivo* patient samples and 1 sample derived from a xenograft model [54].

Only when we analyzed individual patients, we could find the induction of known cell death regulators, such as BIM (*BCL2L11*). Due to the averaging affect when looking for common patterns in populations, these genes were lost, suggesting that each subclass and maybe even each patient might have to be treated as an own entity if one aims to understand the molecular pathway of GC-induced cell death. Similarly, the extent, diversity and magnitude of GC-induced gene regulation seen in individual T-ALL patients is similar to those found in hyperdiploid or ’other’ preB-ALL patients (Supplementary Figure S2), but is lost when looking for common GC-regulated genes in this subset of ALL patients (Figure 2A).

By asking whether the extent of gene regulation by GC correlates with the clinical response, none of the regulated genes showed any significant correlation. The extent of gene regulation, however, did in fact correlate with the GC levels as determined by GBA, demonstrating that the obtained gene expression profiles did reflect the effect of GCs (data not shown). In contrast, the GC-serum levels did not correlate with the clinical response, again indicating that the extent of gene regulation by GCs is not correlated with the clinical response. In summary, comparison of gene expression and regulation by GC in chALL samples did not reveal genes that might explain the drop in blast counts. This might be due to the heterogeneity between and even within subtypes of ALL or it might be due to the fact that GCs regulate many genes which cooperate in cell death induction.

Primary or secondary GC-resistance is caused by mutations in the GR or changes in GR expression levels but no other downstream mechanisms mediating GC resistance have been identified although several genes are known to control GC sensitivity. One possible explanation for this observation is that GCs regulate several different genes involved in metabolism, cell growth, differentiation, etc., to which cells respond with cell cycle arrest and apoptosis. To address whether the combination of GC-regulated genes (’regulators’) in a certain context defined by the expression of other genes (’modulators’) could explain the effect of GC in chALL, we applied elastic net regularization, which led to the definition of gene signatures that corresponded with the clinical outcome.

Among the modulator genes we find CD24, USP28 and GRK5, whose expression levels negatively correlated with the blast response. High expression of CD24, a GPI-anchored receptor involved in B-cell proliferation [55], has been linked to poor prognosis in several tumor entities [56]. USP28 is required to maintain high levels of MYC [57], whose downregulation has been linked to apoptosis in leukemia [58]. Finally GRK5 is a serine/threonine protein kinase that, amongst other targets, phosphorylates p53 to keep it inactive [59]. Thus, our data suggest that low expression levels of CD24, USP28 and GRK5 might not only be prognostic markers but may render leukemia cells susceptible to GCs and possible other forms of chemotherapy. Modulator genes that were found to be positively correlated with the blast response include genes regulating glycosylation and translation, suggesting that cells expressing these genes might depend on extracellular signals and high translation rates, pathways that are known to be targeted by GCs.

In the set of downregulated genes (gene set 1), we find genes involved in metabolism, e.g. glucose transporters, biosynthesis, e.g., ribosome formation and transcription, or mitogenic signaling. While good responders downregulated these genes, poor responders, such as patients with T-ALL, largely failed to downregulate these potentially survival promoting genes. Similarly, the good responders tended to increase gene set 3, which includes genes such as GIMAP2 [60], LXN [61], PCDH9 [62] as well as miRNA mir101-1 [63], which have all been implicated in adversely affecting proliferation in various malignancies.

In summary, our effort to identify transcriptionally regulated genes that can explain the effect of GCs during the first week of treatment of ALL patients, suggests that GCs do not act via regulation of a conserved genes controlling cell death. To improve our understanding of GC-induced cell death, it will be essential to increase the number of cases for each individual subtype of chALL to improve this unique dataset of *in vivo* GC-responses, which will finally help to decipher cell type specific pathways for cell death induction by GCs *in vivo*.

## Data Availability

Raw and preprocessed expression data has been deposited at the Gene Expression Omnibus (Accession number: GSE73578); during review process the data is accessible using the link: http://www.ncbi.nlm.nih.gov/geo/query/acc.cgi?token=sfktcoyghzavrup&acc=GSE73578.

## Declarations

### Ethics approval and consent to participate

Ethics Committee of Innsbruck Medical University (EK1-1193-172/35) with written informed consent obtained from parents or custodians.

## Availability of data

All microarray data are available at:

*http://www.ncbi.nlm.nih.gov/geo/query/acc.cgi?token=sfktcoyghzavrup&acc=GSE73578.*

## Author contribution

TA: performed bioinformatical analysis and generated figures, SS: contributed to writing, DB: contributed to bioinformatical data analysis, AK: performed GBA assay, NSM: devised and supervised machine learning experiments, BM, RC, GM, and RP-G obtained clinical information and were involved in patient treatment, RK: devised, organized and supervised experiments, analysed data and wrote the manuscript, JR: supervised and performed bioinformatical analysis, SG: analysed data and wrote manuscript.

## Acknowledgements

The authors thank Prof. Z. Trajanoski for helpful discussions, K. Götsch, C. Grubbauer, V. Zyka for unpublished data, and M. Brunner, B. Gschirr, A. Kofler for microarray analyses.

